# Spatial versus angular resolution for tractography-assisted planning of deep brain stimulation

**DOI:** 10.1101/19008813

**Authors:** Luka C. Liebrand, Guido A. van Wingen, Frans M. Vos, Damiaan Denys, Matthan W.A. Caan

## Abstract

Given the restricted total scanning time for clinical neuroimaging, it is unclear whether clinical diffusion MRI protocols would benefit more from higher spatial resolution or higher angular resolution. In this work, we investigated the relative benefit of improving spatial or angular resolution in diffusion MRI to separate two parallel running white matter tracts that are targets for deep brain stimulation: the anterior thalamic radiation and the supero-lateral branch of the medial forebrain bundle. Both these tracts are situated in the ventral anterior limb of the internal capsule, and recent studies suggest that targeting a specific tract could improve treatment efficacy. Therefore, we scanned 19 healthy volunteers at 3T and 7T according to three diffusion MRI protocols with respectively standard clinical settings, increased spatial resolution of 1.4 mm, and increased angular resolution (64 additional gradient directions at b=2200s/mm^2^). We performed probabilistic tractography for all protocols and quantified the separability of both tracts. The higher spatial resolution protocol improved separability by 41% with respect to the clinical standard, presumably due to decreased partial voluming. The higher angular resolution protocol resulted in increased apparent tract volumes and overlap, which is disadvantageous for application in precise treatment planning. We thus recommend to increase the spatial resolution for deep brain stimulation planning to 1.4 mm while maintaining angular resolution. This recommendation complements the general advice to aim for high angular resolution to resolve crossing fibers, confirming that the specific application and anatomical considerations are leading in clinical diffusion MRI protocol optimization.

## Introduction

Deep brain stimulation (DBS) is an established treatment for movement disorders and is also used for several treatment-refractory psychiatric disorders, such as obsessive-compulsive disorder (OCD) and major depressive disorder (MDD). DBS for these indications has been successful in multiple targeted brain regions (Alonso et al., 2015), although there is a shift towards understanding DBS as a network effect (Figee et al., 2013), with an increased focus on targeting specific white matter bundles visualized with tractography (Noecker et al., 2017).

The (ventral) anterior limb of the internal capsule (ALIC) is a popular DBS target for psychiatric indications (Tierney, Abd-El-Barr, Stanford, Foote, & Okun, 2014). Earlier work on the neuroanatomy of the ALIC has shown that there is a large amount of inter-subject variance in the white matter organization (Coenen, Panksepp, Hurwitz, Urbach, & Mädler, 2012; Makris et al., 2016; Nanda, Banks, Pathak, & Sheth, 2017). This suggests that accurate subject-specific tractography is required for clinical applications. Tractography studies have pointed towards the supero-lateral medial forebrain bundle (slMFB) (Coenen et al., 2012; Liebrand et al., 2019), and, more recently, to the anterior thalamic radiation (ATR) (Baldermann et al., 2019), as the preferred DBS target in the ALIC. The ATR connects the anterior thalamus with the prefrontal cortex, while the slMFB constitutes the rostral part of the cortico-pontine connection between the ventral tegmental area and prefrontal cortex (Coenen et al., 2012). When passing through the ALIC, these bundles are oriented in approximately the anterior-posterior direction with, typically, a respective medial-lateral organization.

In order to refine tractography-assisted DBS targeting, and evaluate the relative benefits of slMFB versus ATR stimulation in the ALIC, more prospective tractography studies are necessary. DBS targeting requires high precision, and in our experience slight alterations in placement may affect the treatment outcome (Liebrand et al., 2019). Therefore, it is important to optimize diffusion-weighted imaging (DWI) acquisitions and develop robust tractography pipelines for these studies. Current clinical applications of tractography are often based on the diffusion tensor imaging (DTI) model (Petersen et al., 2016), which enables reproducible reconstruction of the major white matter tracts, despite the low spatial and angular resolution of the DWI’s (Wakana et al., 2007). However, the DTI model cannot account for more complex fiber configurations, such as fibers crossing or touching, which may occur in up to 90% of white matter voxels (Jeurissen, Leemans, Tournier, Jones, & Sijbers, 2013). It is therefore suggested to acquire DWI data at a higher angular resolution and with multiple b-values, in order to use more sophisticated models that can resolve multiple fiber populations within each voxel (Caan et al., 2010).

Here, we focus on optimizing the acquisition for distinguishing the slMFB and ATR in the ALIC with tractography for use in a clinical setting. It can be reasoned that resolving parallel fibers in the ALIC does not require high angular resolution compared to crossing fiber configurations, so that, as long as the angular resolution (i.e. diffusion sensitivity) is sufficient to detect tract orientations, clinical scanning time can be spent on improving spatial resolution (i.e. voxel size). For this reason, we investigate the tradeoff between angular and spatial resolution for reconstructing and distinguishing between both tracts by comparing a standard clinical 3T protocol to respectively high angular (3T) and high spatial (7T) resolution protocols. Tract reconstructions for each protocol will be evaluated based on their capability to resolve the slMFB and ATR, to choose the optimal tradeoff for specific targeting of either bundle.

## Materials and Methods

### Participants

Nineteen healthy volunteers were included in this study approved by the Institutional Review Board after giving written informed consent. After screening for MR contraindications, all participants were scanned in 3T and 7T scanners, with both scanning sessions taking place on the same day in the period between August and October, 2017. Structural (T1 and T2-weighted) scans were made at 3T to allow for comparison between the diffusion protocols in individual structural space.

### Scan protocols

Data were acquired on Philips Ingenia 3T and Achieva 7T scanners (Philips Medical Systems, Best, Netherlands) equipped with 32-channel phased-array head coils (Nova Medical, Wilmington, MA). A complete overview of the applied diffusion MRI scan parameters is given in Table 1. Three diffusion-weighted scans were made: a standard scan with parameters similar to clinical protocol of around 3 minutes, and two high resolution scans with a duration of approximately 10 minutes, which is a timescale that is usable in the clinic. One high resolution scan included more diffusion orientation measurements (higher angular resolution), and the other had a higher spatial resolution, respectively. We will refer to these as the HARDI and HSRDI scans.

**Table 1.** Overview of acquisition parameters for the three sequences. (HARDI: high angular resolution diffusion imaging, HSRDI: high spatial resolution diffusion imaging, FOV: field-of-view)

|  | Standard | HARDI | HSRDI |
| --- | --- | --- | --- |
| Field strength (T) | 3 |  | 7 |
| Field of view (mm<sup>3</sup>) | 224 x 224 x 135 |  | 140 x 179 x 51 |
| Voxel size (mm<sup>3</sup>) | 2.3 x 2.3 x 2.3 |  | 1.4 x 1.4 x 1.4<br>(10% slice gap) |
| Acceleration | SENSE = 1.5<br>Multiband = 2 |  | SENSE = 2.7<br>80% FOV in phase<br>encoding direction |
| Half-scan | 0.7 |  | 0.7 |
| Repetition time (ms) | 5363 |  | 3038 |
| Echo time (ms) | 99 |  | 71 |
| b-values (s/mm<sup>2</sup>) | 32 x b = 1000 | 32 x b = 1000<br>64x b = 2200 | 32 x b = 1000 |

| Signal averages | 1 |  | 3 |
| --- | --- | --- | --- |
| Scan time | 3:07 min | 9:44 min | 10:13 min |

The standard scan was made at 3T with an isotropic 2.3 mm voxel size, and contained 32 volumes with non-collinear diffusion directions and a diffusion weighting of b=1000 s/mm^2^ and a single b0-reference volume. The HARDI scan extended the standard scan with an additional shell of 64 directions at a higher diffusion weighting value (b=2200 s/mm^2^) to investigate the relative importance of angular resolution. The scanning was further expedited by applying a multiband factor of 2.

The HSRDI scan was made at 7T at a higher spatial resolution of 1.4 mm^3^, and contained the same number of diffusion-weighted volumes as the standard scan (i.e., 32x b=1000 s/mm^2^, 1x b=0), acquired without multiband acceleration. The field-of-view was slightly reduced in the z-direction (feet-head) to keep scanning time in line with the 3T scans. In order to minimize the distortions that come with longer readout times when scanning at higher spatial resolution, the field-of-view (FOV) was reduced in the phase encoding direction, with additional outer volume suppression to prevent aliasing (Gallichan, 2018; Heidemann, Anwander, Feiweier, Knösche, & Turner, 2012). In order to correct for distortions during preprocessing, b0-reference volumes with opposite phase encoding polarity were also acquired at 3T and 7T.

### Diffusion (pre)processing

The raw diffusion data were first converted to nifti before noise filtering with a PCA-based filter (Veraart et al., 2016), as implemented in MRtrix3 (https://mrtrix.readthedocs.io/). Distortion, motion and eddy current correction was subsequently performed with FSL’s (FMRIB’s Software Library, version 5.0.10; https://fsl.fmrib.ox.ac.uk/fsl/) TOPUP and *eddy_cuda* (gpu version of *eddy*) tools (Andersson, Graham, Zsoldos, & Sotiropoulos, 2016; Andersson & Sotiropoulos, 2015). Finally, the data was corrected for Gibbs’ ringing artifacts with an in-house developed Matlab (The Mathworks, Natick, MA; version R2017a) script. The preprocessed diffusion data were fit with a diffusion tensor model for visual inspection (see Figure 1), and with FSL’s Bedpostx, a ball-and-stick model capable of modelling multiple diffusion orientations per voxel (Behrens, Berg, Jbabdi, Rushworth, & Woolrich, 2007), for tractography analysis.

**Figure 1.**
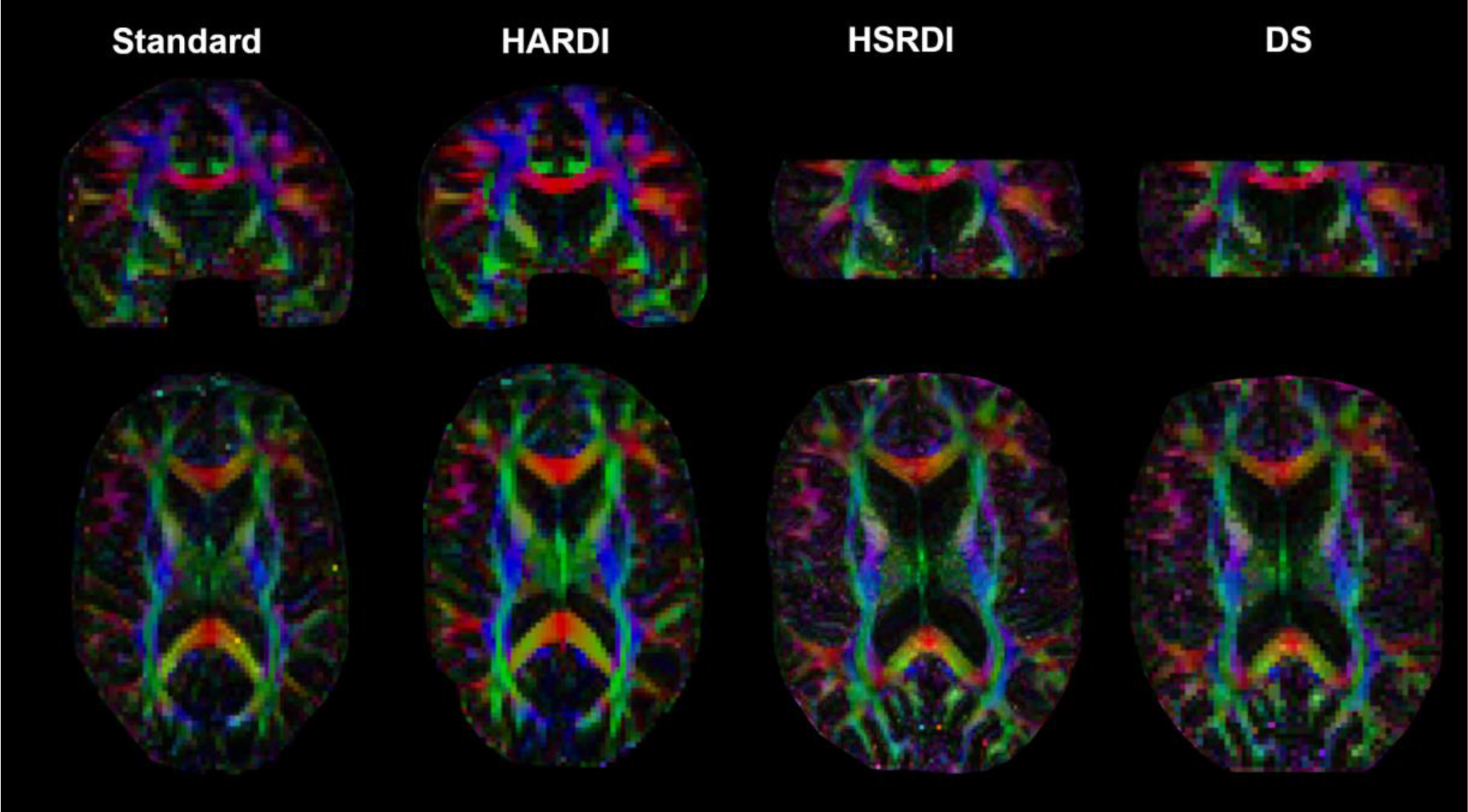
Comparison of coloured fractional anisotropy maps between the four datasets (standard, HARDI, HSRDI, and DS, respectively) in native diffusion space for one participant. Note that the slices are not exactly at the same location and orientation because of differences in acquisition parameters and participant position in the scanner. The difference between the HSRDI and the HARDI scans is immediately apparent, whereas the differences between the standard and HARDI scans are more subtle. The HSRDI scan shows more anatomical detail and suffers less from partial voluming, resulting in increased distinction between separate directions. The HARDI is less noisy and looks cleaner than the baseline scan due to the additional volumes. The DS dataset appears very similar to the standard acquisition.

### Down-sampled dataset

To ensure that eventual differences between the HSRDI dataset and standard are not caused by field strength, we additionally performed tractography in a down-sampled 7T dataset (hereafter ‘DS’). The HSRDI dataset was down-sampled to match the spatial resolution of the other datasets after denoising and Gibbs ringing correction, since the correction tools recommend not using interpolated data. Further processing was identical for all datasets.

### Tractography analysis

Rigid co-registrations between diffusion and and structural space were performed with SPM (statistical parametric mapping, version 12; https://www.fil.ion.ucl.ac.uk/spm/) based on the fractional anisotropy (FA) maps and T1-scans, and visually inspected. Tractography seeds for the ATR and slMFB were drawn in the anterior thalamic nucleus and ventral tegmental area, respectively, with an additional waypoint in the anterior limb of the internal capsule, according to Coenen et al. (2012). The seeds were subsequently transformed to diffusion space. Probabilisitc tractography was performed with FSL’s Probtrackx between the seed and waypoint for both bundles bilaterally (default settings). The tract reconstructions were transformed to structural space and resliced to 1 mm isotropic resolution, after which they were visually assessed. See Figure 2 for a tractography example of the ATR and slMFB.

**Figure 2.**
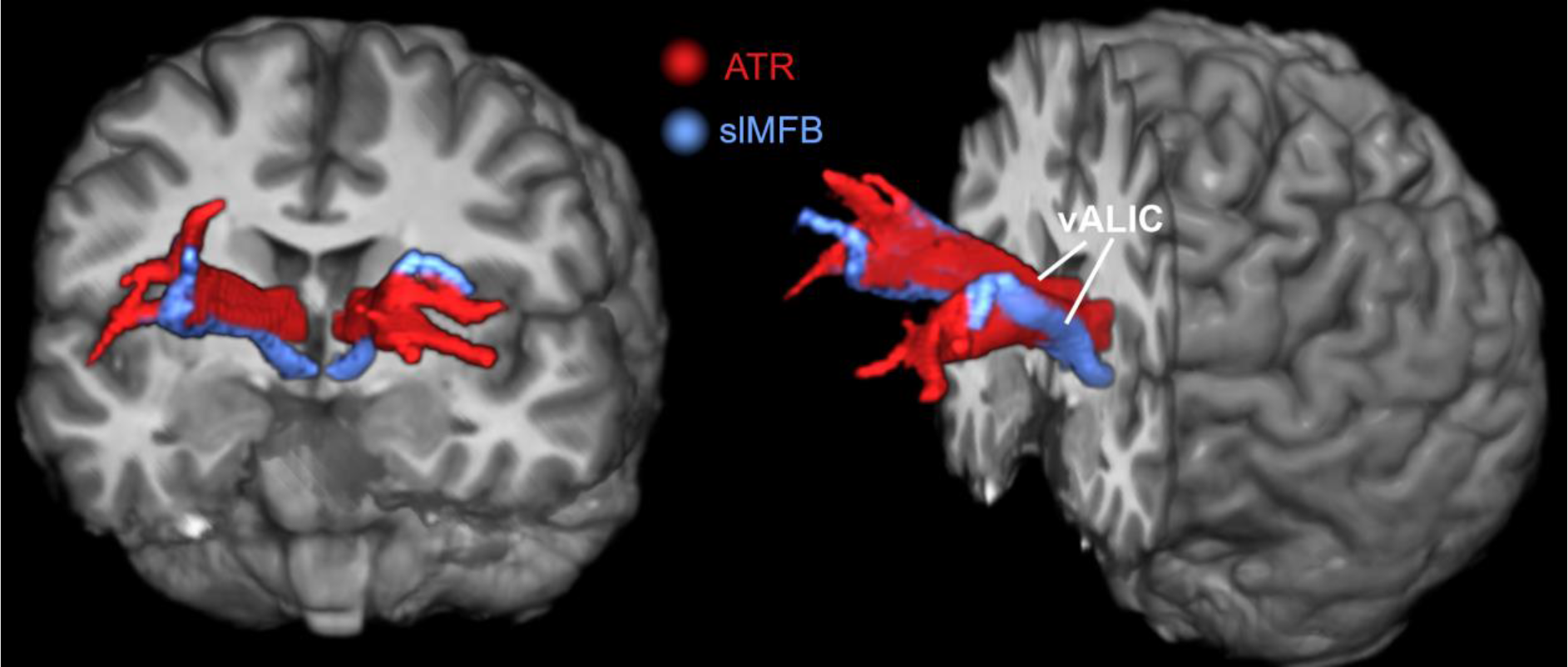
Three-dimensional tractography overview of the ATR (red) and slMFB (blue) on top of a structural MR image. In the ventral ALIC (as indicated by the lines), the slMFB runs lateral to the ATR. Towards the prefrontal cortex, the bundles are more intermixed, as they branch out and connect to adjacent prefrontal structures. The diffusion data in this figure were acquired to the HSRDI protocol.

### Comparison of tract probability profiles and statistical analysis

We compared the separability of the ATR and slMFB in the anterior limb of the internal capsule per sequence. In order to preserve the spatial information of each individual, we performed the tractography analysis in subject space. To compare tract probability profiles across subjects and DWI sequences, we quantified their separability. This was done in the coronal plane, which is oriented approximately perpendicular to the anterior-posterior axis orientations of the ATR and slMFB (Coenen et al., 2012). For each individual, the target area for DBS was identified in the vALIC on the T1-weighted scan. Along the line on the medial-lateral axis through this target area, normalized track probability profiles of the ATR and slMFB were computed, as shown in Figure 3. We calculated the mean standardized difference (MSD) as a measure of bundle separability, which is defined as the distance between the two peaks divided by the average standard deviation (full width at half max, FWHM). Five adjacent lines per hemisphere were averaged to improve robustness of this measure. Finally, we performed a repeated-measures analysis of variance (ANOVA) in Matlab (*fitrm*), with Greenhouse Geisser non-sphericity correction when necessary, to assess possible differences between the three scans (standard, HARDI, and HSRDI). In case of significant differences, we calculated pairwise differences post-hoc with Bonferroni correction for multiple comparisons (*ttest*).

**Figure 3.**
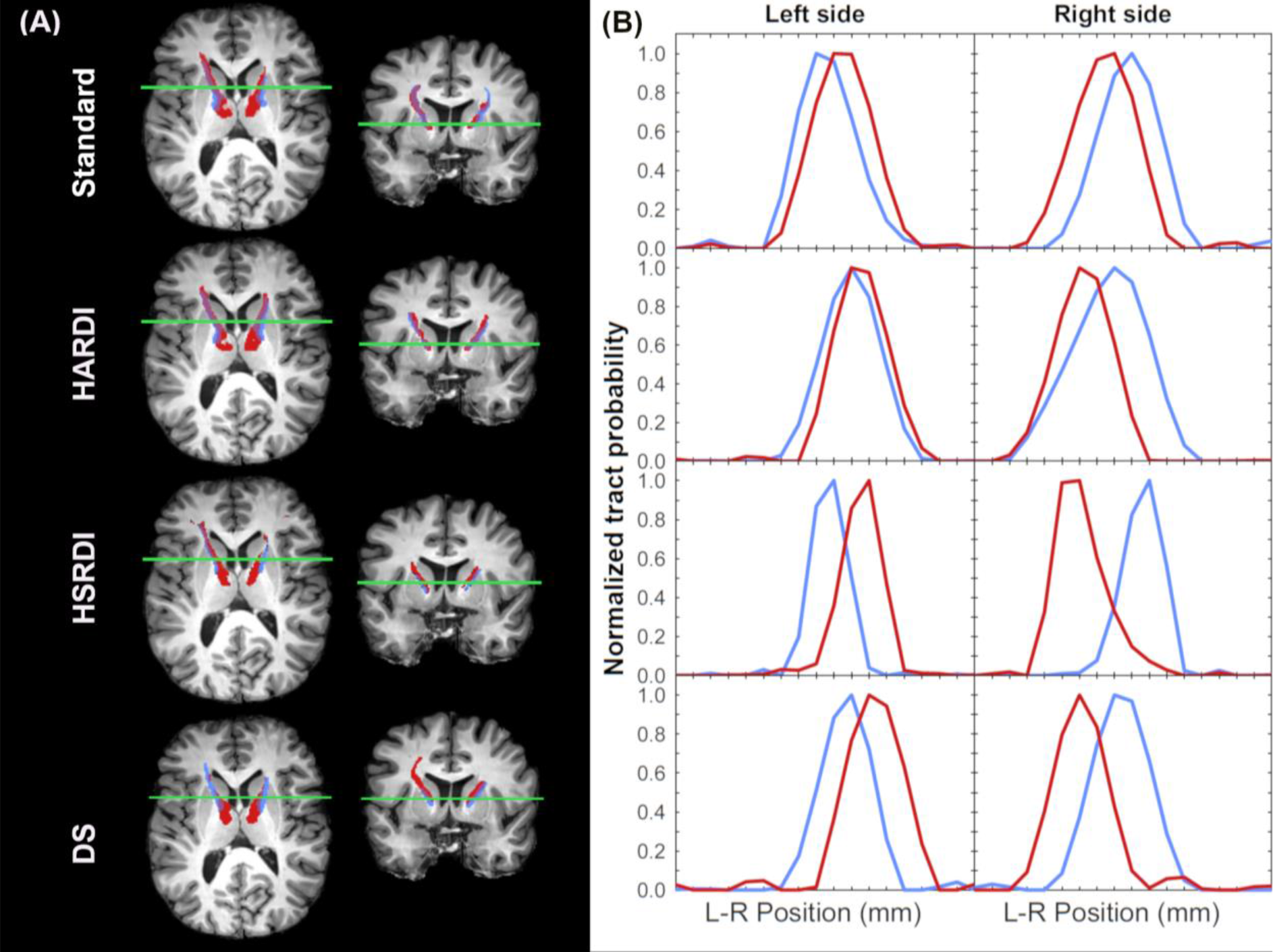
(A) Co-registered tractography results of the ATR (red) and slMFB (blue) for all four datasets (rows) and (B) corresponding tract probability profiles for one subject. Since tract probabilities have arbitrary scaling, there may be slight differences in bundle appearance due to differences in thresholding. Tractography results for the standard, HARDI, and DS sequences are quite similar in shape and size, with the HARDI tracts being slightly more voluminous than the baseline. In contrast, the HSRDI tracts appear much narrower and seem to fit the white matter anatomy better. This is also reflected in the tract probability profiles, which show that the peak-to-peak distance does not vary by much between the sequences, although there is a visual difference between the bundle widths.

## Results

Data collection of two participants could not be completed at 3T, as a result of technical issues; furthermore, one participant withdrew and was not scanned at 7T. These participants were excluded from the analysis to yield 16 complete datasets. We were able to reconstruct the slMFB and ATR bilaterally for all DWI sequences in every participant except one, for whom the inferior part of the slMFB was outside of the HSRDI scan’s field of view. In order to keep the groups balanced, we excluded the other scans from this participant from further analysis. Therefore, the final analysis consisted of 45 DWI scans of fifteen participants (mean age = 32 ±8 years, 5 females). An overview of all tractography results of can be found in Supplementary Figure 1.

Figure 4 shows the distribution of MSD’s between the ATR and slMFB stratified by scan protocol. It can be seen that the HSRDI protocol yielded a higher average MSD than the standard protocol (41% average increase), whereas the HARDI protocol yielded a lower average MSD than the standard protocol (29% average decrease). The DS dataset performed similar to the standard dataset in all aspects. A repeated-measures ANOVA showed that there was a significant difference in MSD between groups (F(3,42) = 19.4, p = 10^−5^). Further assessment of pairwise differences (corrected for multiple comparisons) demonstrated that the MSD of the HSRDI sequence was significantly larger than the other datasets (HSRDI-vs-standard t(14) = 3.6, p = 0.01; HSRDI-vs-HARDI: t(14) = 6.2, p = 10^−4^; HSRDI-vs-DS: t(14) = −8.3, p < 10^−5^). The MSD was also significantly higher for the standard dataset compared to the HARDI scan (HARDI-vs-standard: t(14) = 4.2, p 0.01).

**Figure 4.**
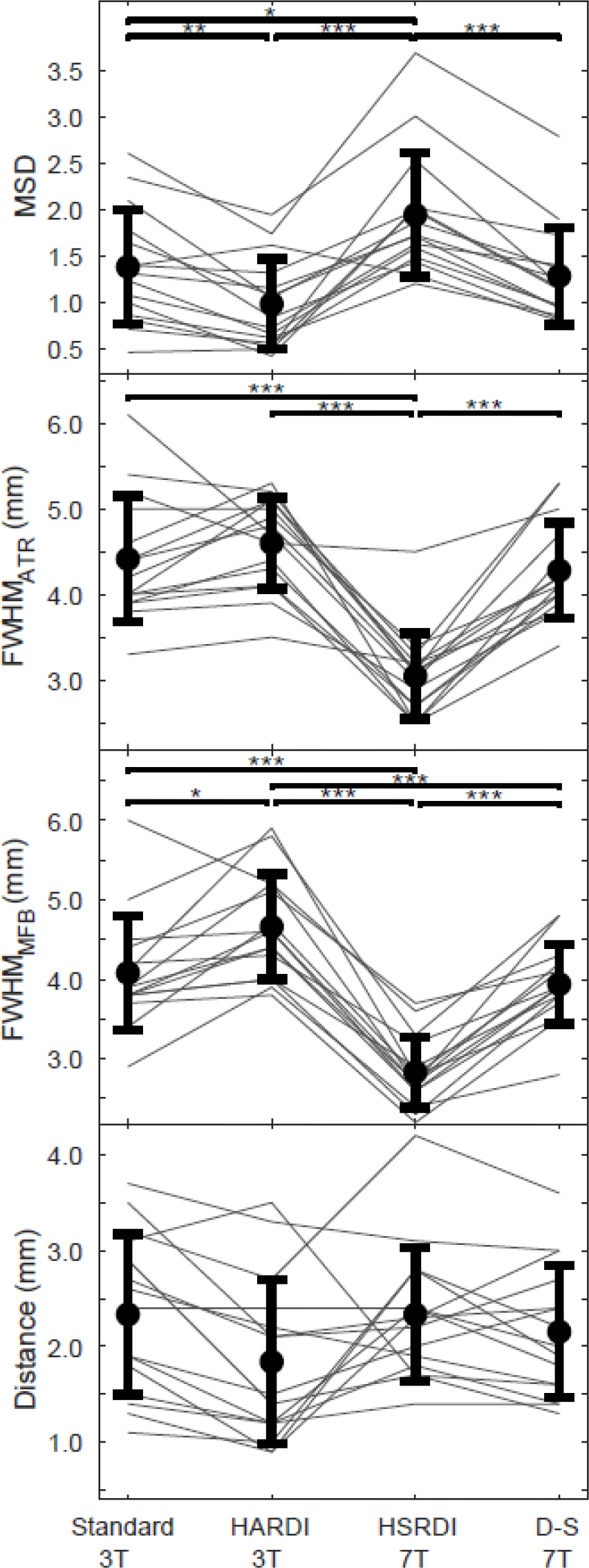
Comparison of the mean standardized difference (MSD), full-width-at-half-max (FWHM) and the distance between ATR and slMFB for all datasets. In all four panels, errorbars indicate the mean and standard deviations, with the separate data points for each subject connected by thinner lines. The top panel shows that the HSRDI sequence yields a higher MSD than the other three datasets. The HARDI scan results in a decreased MSD compared to the standard and DS images, although the gray lines show that this is not the case for every subject. In the middle two panels, the FWHM of respectively the ATR and slMFB are shown, showing that these are smaller for the HSRDI compared to the other sequences, and very similar between both bundles. There is no significant difference between the average distance from ATR to slMFB between sequences (bottom panel). Bars are only shown for significant differences, with significance levels indicated by the asterisks: * for p<0.05; ** for p<0.01; *** for p<0.001.

To investigate whether the difference in MSD between sequences was caused by alterations in peak-to-peak distances between the bundles, or by differences in the width of the tract profiles (i.e. the FWHM), these parameters were also plotted in Figure 4. Repeated-measures ANOVAs showed significant differences between the FWHM of the ATR (F(3,42) = 39.2, p < 10^−11^) and MFB (F(3,42) = 59.7, p < 10^−14^), respectively. Post-hoc assessment of pairwise differences (corrected for multiple comparisons) found that the HSRDI sequence produced a significantly lower FWHM compared to the standard and HARDI sequences for the ATR (HSRDI-vs-standard: t(14) = −9.7, p < 10^−6^; HSRDI-vs-HARDI: t(14) = −13.2, p < 10^−8^) and slMFB (HSRDI-vs-standard: t(14) = 9.7, p < 10^−6^; HSRDI-vs-HARDI: t(14) = 13.2, p < 10^−7^; HSRDI-vs-DS: t(14) = 9.1, p < 10^−5^). There were also smaller, but still significant, differences in the FWHM of the standard and DS datasets on one hand and HARDI on the other for the slMFB (Standard-vs-HARDI: t(14) = −3.5, p = 0.01; DS-vs-HARDI: t(14) = 5.3, p < 10^−5^). In contrast, the peak-to-peak distances were very similar between sequences and did not show any significant differences.

## Discussion

The aim of this study was to optimize separability of the ATR and slMFB in the ALIC for specific targeting of either bundle in prospective DBS studies. We investigated the relative importance of angular and spatial resolution in diffusion MRI with limited scan time, as is common in clinical practice. DWI data were acquired according to one standard and two extended scan protocols with, respectively, increased angular and spatial resolution from a group of healthy volunteers. We performed probabilistic tractography to reconstruct the ATR and slMFB in all four datasets (i.e. three acquisitions and one down-sampled dataset). Finally, the mean standardized difference (MSD) was calculated as a measure of the separability of the ATR and slMFB. The increased spatial resolution (HSRDI) sequence yielded a significantly larger MSD than the other three datasets, suggesting that it was better able to separate the two bundles.

The 7T (HRSDI) sequence with increased spatial resolution had better separability due to the smaller apparent cross-sectional area of the bundles, as there was no statistically significant difference in the peak-to-peak distances between the ATR and slMFB of all four datasets. Because of reduced partial voluming at higher spatial resolution at 7T in the plane perpendicular to the bundle trajectory, the overlap between ATR and slMFB could be reduced, thereby improving the separation of the tracts. There were no complex fiber crossings that needed to be resolved in the ATR and slMFB bundles using higher b-values data. It follows that the clinically used b-value of 1000 s/mm^2^ was sufficient for accurate tract reconstruction and that increasing spatial resolution benefitted tract reconstruction.

The HARDI sequence actually yielded a small but significant decrease of the MSD compared to the standard sequence. This can be attributed to the significant increase in bundle cross-sectional area of the slMFB in the HARDI protocol. We hypothesize that the apparent increase in tract cross-sectional area originates from the improved sensitivity to diffusion provided by the additional diffusion-shell. This is supported by our observation (data not shown) that the anisotropic volume fraction of the diffusion model used for tractography increased throughout the white matter, when adding the additional shell with a higher b-value. Effectively, this leads to wider tract outlines. Nevertheless, we have not attempted to quantify the volume of white matter tracts, since quantification of white matter volume and integrity with tractography is inherently limited (Jeurissen, Descoteaux, Mori, & Leemans, 2019; Jones & Cercignani, 2010). Still, it remains possible that the apparent tract cross-sectional area at higher spatial resolutions is more reflective of the actual bundle cross-sectional area than at lower spatial resolutions.

The debate about optimal angular and spatial resolution in DWI acquisitions for tractography is still ongoing. Animal experiments suggest that angular and spatial resolution must be balanced (Calabrese et al., 2014). A later study in the human brain recommended to acquire DWI data at high angular resolution, since scans at a high angular resolution (obtained by applying strong diffusion-weighting gradients along many gradient directions) may help to resolve multiple fiber populations within each voxel (Vos et al., 2016). Conversely, data acquired at a high spatial resolution may increase the uniformity of tissue structures within voxels by decreasing the voxel volume, revealing structure previously unseen (Steele et al., 2017). It is feasible to increase both angular and spatial resolutions simultaneously, but this leads to increasingly longer scan times that may be unsuitable for patient studies and clinical use. Therefore, the choice of angular and spatial resolution may well depend on the white matter bundles being studied (Calabrese et al., 2014).

While high-resolution 7T clinical applications of diffusion MRI exist for DBS (Patriat et al., 2018; Plantinga et al., 2018), 7T hardware availability is limited. With ongoing developments in acceleration techniques such as multiband, there is potential for higher spatial resolution acquisitions at 3T. Moreover, data processing packages have become better able to deal with distorted (Andersson & Sotiropoulos, 2015) and noisy data (Veraart et al., 2016), allowing to more readily push for higher spatial resolutions in diffusion acquisitions. Therefore, it should be possible to acquire higher spatial resolution DWI data for clinical use at 3T.

Challenges in establishing the tracts’ volumes have the added consequence that absolute distance measurements in tractography suffer from uncertainty (Jones & Cercignani, 2010; Jones, Knösche, & Turner, 2013). Nevertheless, the relative differences in FWHM and the distance between the probability profiles give an indication of how well we can distinguish between the ATR and slMFB. The locations of the peak probabilities of both bundles were consistent between sequences, which is supportive to the consistency of the ATR and slMFB tract reconstructions. Since the locations of the peak probabilities were consistent, there were no significant differences in the distances between tracts per sequence.

Current clinical practice in tractography-assisted surgery often relies on deterministic tractography, as opposed to probabilistic tractography (Petersen et al., 2016). In deterministic tractography, there is only one solution for fitting a tractography algorithm to the underlying diffusion MRI data. As a result, it may appear that there is a sharp transition between two bundles, where in reality the bundles are likely to overlap (Coizet et al., 2017). Therefore, we calculated the MSD between two bundles from probabilistic tractography, anticipating that it will provide a more reliable indication of the suitability of the data for tract delineation.

### Limitations

There are some limitations to this study. Firstly, the data were acquired at two different field strengths hindering attribution of the results to spatial or angular resolution alone. Tissue properties may differ with field strength, such as decreased T2-relaxation times at 7T that require shorter echo times. Although other parameters than the echo time also varied between the 3T and 7T sequences, we do not believe that this invalidated our comparison, as it is arguably better to compare sequences with optimized parameters with each other rather than to copy all parameters between sequences. Importantly, the diffusion process can be expected to be unaffected by magnetic field strength. In addition to above arguments, and to fully rule out the effect of field strength on the results, we down-sampled the 7T dataset and compared it to 3T data at the same resolution. This comparison showed a remarkable agreement in tractography results, suggesting that our results did not depend on field strength.

Secondly, the high angular resolution 3T data was not optimized for spatial resolution, since it was our aim to compare data acquired with clinically used parameters to respectively higher angular and spatial resolution alternatives. Higher spatial resolutions in high angular resolution diffusion imaging have been used previously. For instance, data were acquired for the human connectome project (HCP) at a spatial resolution of 1.5 mm isotropic, even for very high b-values up to b = 10,000 s/mm^2^ (Setsompop et al., 2013). However, the combination of higher angular and spatial resolutions require specialist scanner hardware (e.g., stronger gradients and coils with a large number of channels), which makes these scan parameters difficult to attain in a clinical setting. In this light, the use of default clinical parameters and clinically attainable scan times in our study facilitates clinical application.

A third limitation lies in the trade-off between angular and spatial resolution. Many different parameter combinations exist that increase the angular resolution of a diffusion MRI scan. We limited ourselves to investigating only one protocol with improved angular resolution. This protocol’s parameters are common in high angular resolution diffusion imaging, albeit with a lower b-value than the reported b = 3000 s/mm^2^ for optimal angular resolution (Tournier, Calamante, & Connelly, 2013). Finally, we have no direct comparison with histological data which could serve as ground truth in determining the bundles’ true dimensions. Despite these limitations, within the limited timeframe available to clinical diffusion MRI scanning, and considering the parallel tract orientations within the vALIC, it seems more advantageous to increase the spatial resolution than the angular resolution for parallel running tract configurations. At the same time, it is likely that increasing spatial resolution at the cost of angular resolution is detrimental in crossing tract configurations, since these may require a higher angular resolution to be resolved. Thus, we limit our recommendation of focusing on spatial resolution to resolving parallel tracts for DBS planning.

### Relevance

This study is relevant to precise delineation of parallel running tracts, such as needed for tractography-assisted targeting in DBS in the ALIC. These (relatively) simple parallel tract configurations have received little attention, while it is our experience that even small differences, on a millimeter scale, in DBS localization can alter treatment efficacy (Liebrand et al., 2019). Additionally, more precise and selective stimulation may potentially decrease side effects and prolong battery life. Especially with the advent of so-called directional, or, steering DBS electrodes (Steigerwald, Matthies, & Volkmann, 2019), which are to an extent able to focus the generated electric field to one side, there is an increased potential of delivering stimulation to precisely defined neuroanatomical structures. Therefore, we aim to implement a higher spatial resolution diffusion MRI scan for pre-surgical vALIC DBS planning for clinical practice.

## Conclusions

We conclude that, within a limited amount of scanning time, it is more beneficial to increase the spatial than the angular resolution to be able to precisely discern the slMFB and ATR in the vALIC, as is required for tract-specific DBS. This is primarily caused by the smaller cross-sectional area of the tracts found at higher spatial resolution, leading to decreased overlap. Dictated by local anatomy, our protocol advice deviates from general recommendations to aim for high angular resolution to resolve crossing fibers. Our work thereby allows for increased precision of tract-specific DBS targeting in the vALIC by increasing the spatial resolution of the diffusion MRI scans. Furthermore, it encourages researchers and clinicians to optimize the scanning protocol to the specific anatomy and application at hand.

## Data Availability

We are not allowed to share data online, since complete anonymization is impossible and the data can be traced back to individual participants.

## Acknowledgements

This study was supported by an Innovation Impulse grant (#2015.011) from the Academic Medical Center. Additionally, the study was financially supported as part of the STW Perspectief programme Population Imaging Genetics (ImaGene) by the Dutch Technology Foundation (STW), project 12722.

---

**Supplementary Figure 1.**
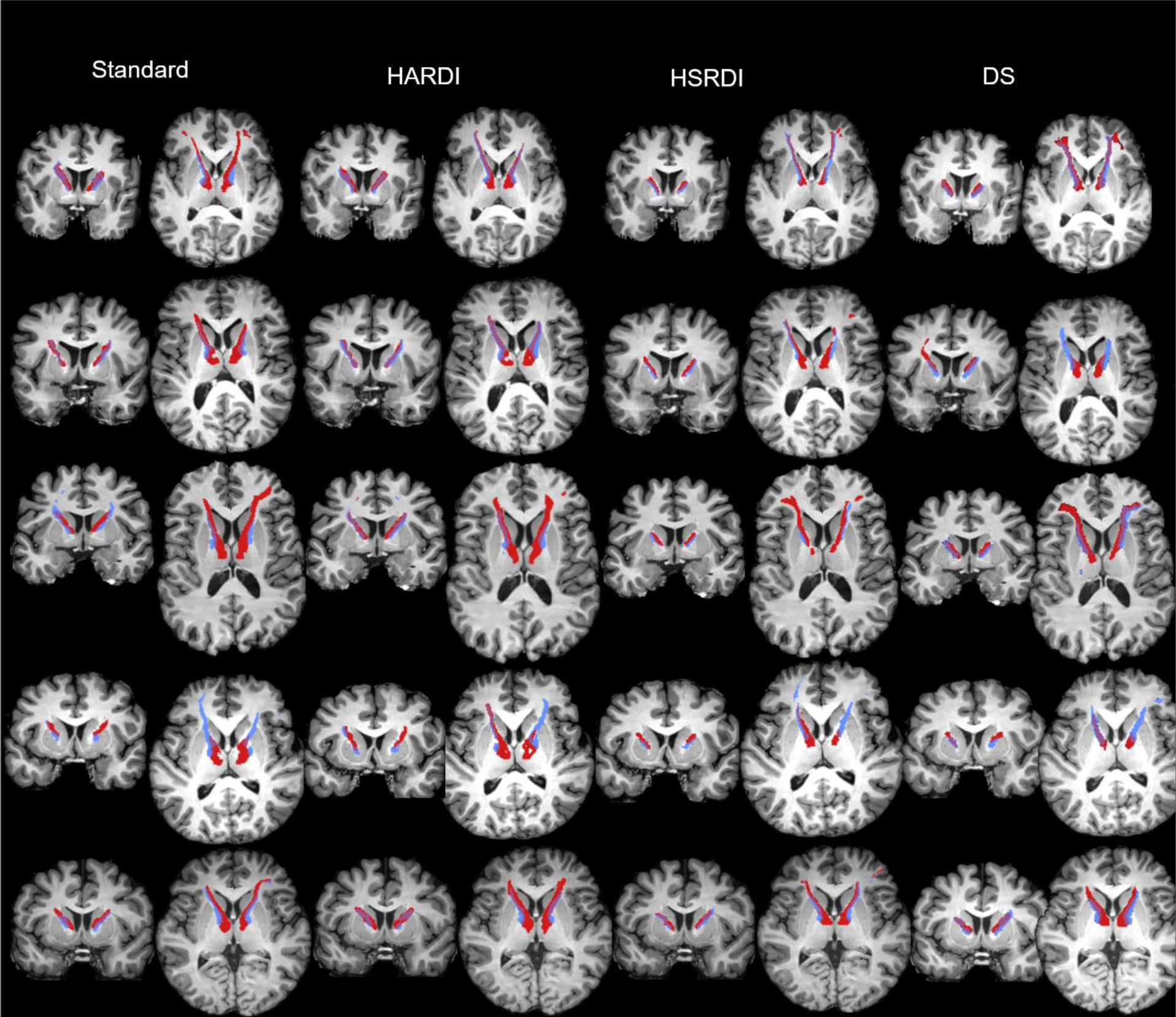
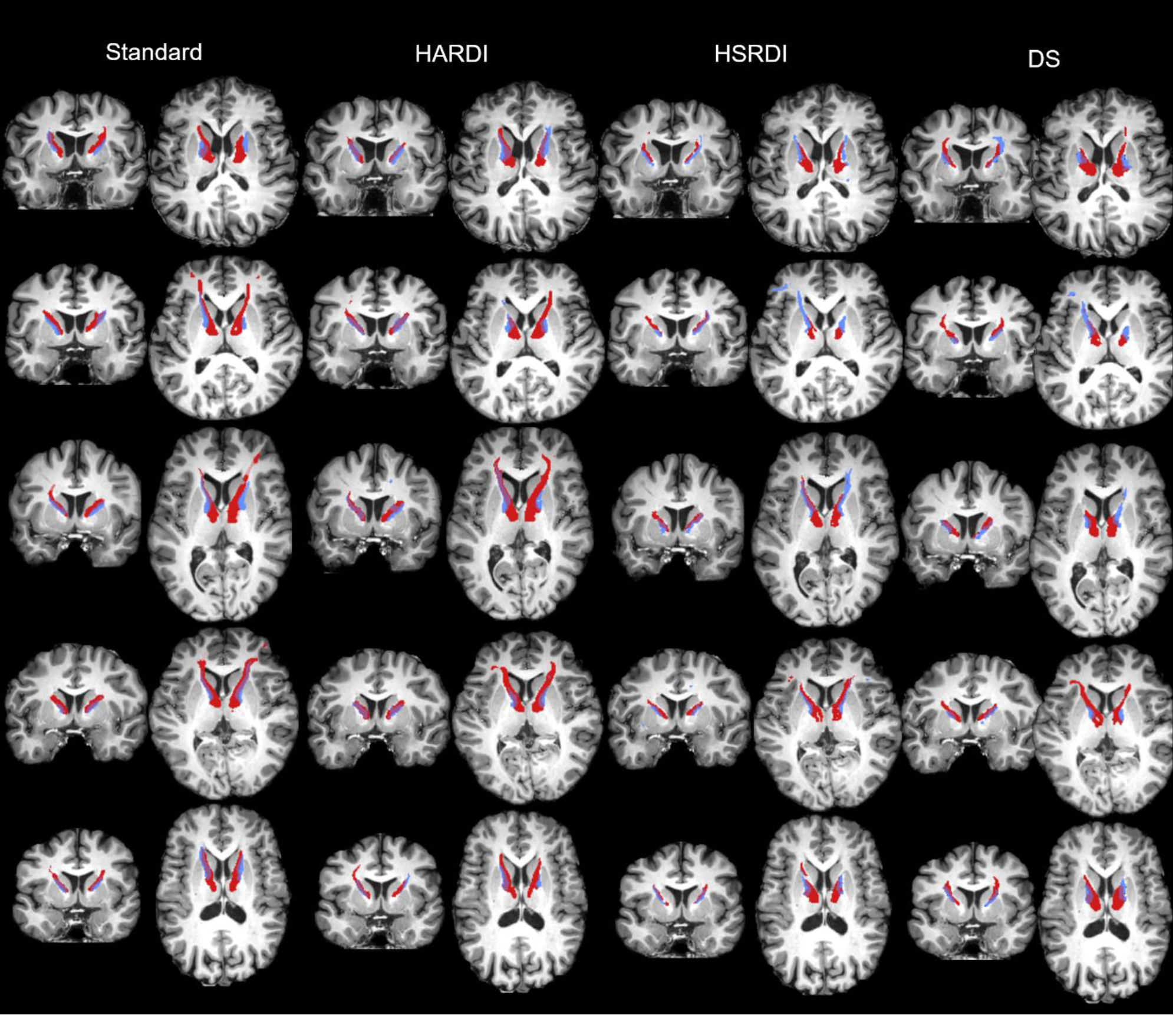
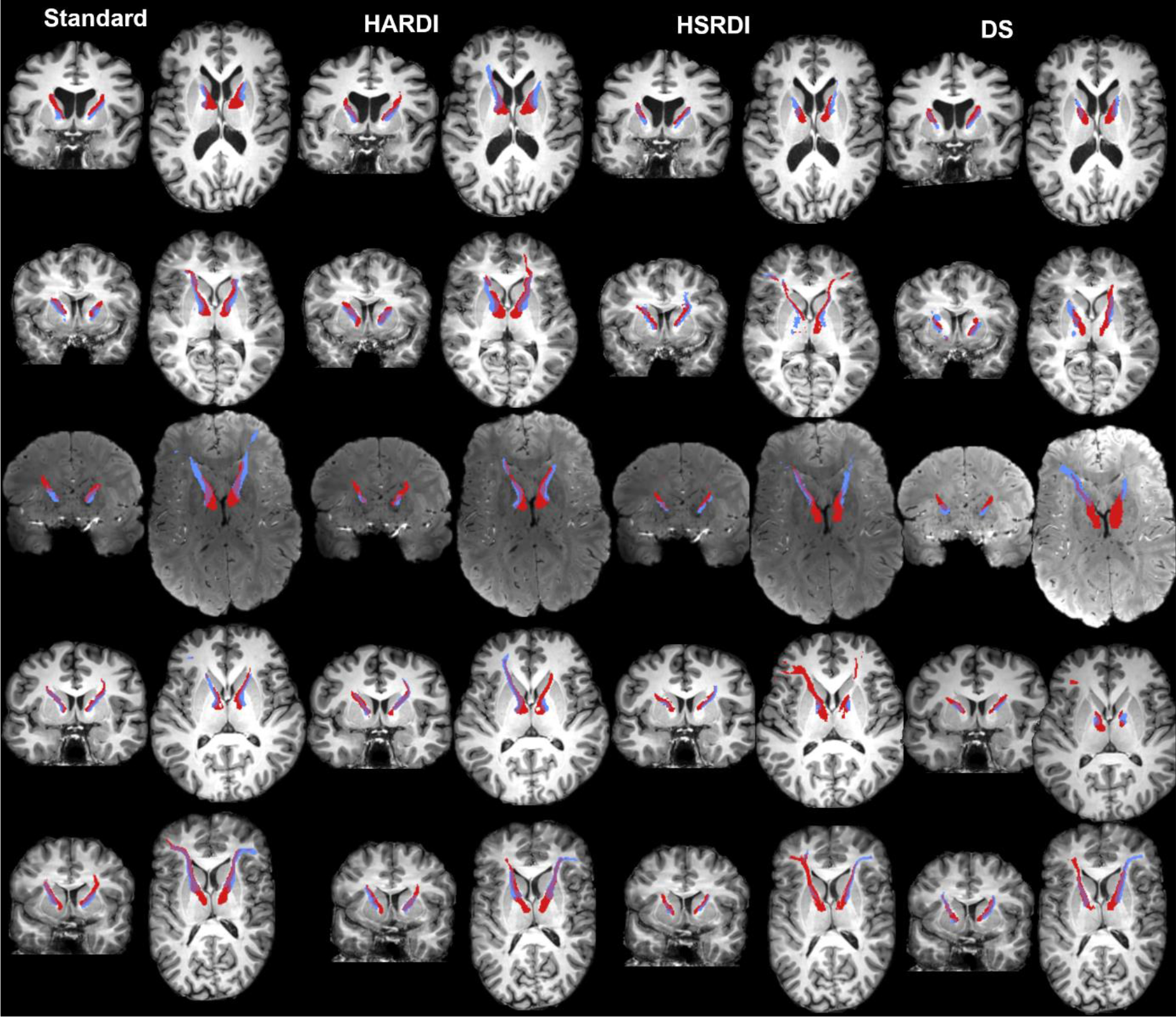
Axial and coronal tractography overviews of the complete sample (N=15) included in this study in individual anatomical space, for all four datasets (standard, HARDI, HSRDI, and DS, respectively). All participants, except one, had a T1-scan available. The colour coding corresponds to the other figures in this paper: the anterior thalamic radiation in red, the supero-lateral branch of the medial forebrain bundle in blue. (HARDI: high angular resolution diffusion imaging; HSRDI: high spatial resolution diffusion imaging; DS: down-sampled)

